# Cardiovascular risk management in adults with diagnosed diabetes in Mexico from 2016–2023: A retrospective analysis of nationally representative surveys

**DOI:** 10.1101/2024.09.18.24313926

**Authors:** Daniel Ramírez-García, Jerónimo Perezalonso-Espinosa, Padme Nailea Méndez-Labra, Carlos A. Fermín-Martínez, Juan Pablo Díaz-Sánchez, César Daniel Paz-Cabrera, Arsenio Vargas-Vázquez, Miriam Teresa López-Teros, David Flood, Jennifer Manne-Goehler, Neftali Eduardo Antonio-Villa, Goodarz Danaei, Jacqueline A. Seiglie, Omar Yaxmehen Bello-Chavolla

## Abstract

**BACKGROUND:** Effective cardiovascular disease (CVD) risk management is a cornerstone of optimal diabetes care. Here, we estimated the prevalence and determinants of CVD risk factor control amongst individuals with diagnosed diabetes in Mexico.

**METHODS:** We analyzed data from individuals with diagnosed diabetes ≥20 years from the 2016-2023 Mexican National Health and Nutrition Surveys. We estimated the prevalence of glycemic, blood pressure (BP), non-current smoking, and combined CVD risk factor control. LDL-C control was assessed using SCORE2-Diabetes risk categories. We estimated the prevalence of BP-lowering, cholesterol-lowering, and glucose-lowering medication use, and explored determinants of control achievement using logistic regression.

**RESULTS:** We analyzed data representing 43.2 million adults with diagnosed diabetes during 2016-2023. In 2023, glycemic control was 29% (95%CI 21%-38%), BP control 22.9% (95%CI 14%-31%), and non-current smoking 89% (95%CI 81%–96%). The proportion of people classified as high or very-high CVD risk increased from 59.8% (95%CI 52.1%-67.0%) in 2016 to 68.4% (95%CI 55.6%-78.9%) in 2023, representing ∼5.1 million adults. LDL-C control prevalence increased from 2.8% (95%CI 1.2%-4.4%) in 2016 to 6.6% (95%CI 1.9%-11.2%) in 2023. Combined risk factor control achievement was low primarily due to suboptimal LDL-C control, despite high medication use; this was more likely achieved in females, younger individuals, and those with college education or living in states with higher socioeconomic position.

**CONCLUSIONS:** Despite increasing CVD risk during this period, comprehensive glycemic and CVD risk factor management for adults with diabetes in Mexico remains suboptimal. Our findings highlight the need for strategies to address gaps in CVD risk management to reduce premature mortality in this population.

**Lay summary:** This study examined how well adults with diabetes in Mexico are controlling key risk factors for cardiovascular disease, such as blood glucose levels, blood pressure, smoking, and cholesterol levels. Authors used nationally representative surveys from 2016-2023 analyzing data which represents over 43.2 million adults living with diagnosed diabetes.

- In 2023, fewer than one-third of individuals with diabetes had adequate blood glucose control, and fewer than one in ten met recommended cholesterol targets, despite high rates of medication use. Cardiovascular risk increased for those at the highest risk by nearly 9% between 2016 and 2023.
- Better control of these risk factors was more common among women, younger individuals, those with higher education, and those living in more socioeconomically advantaged areas.

**GRAPHICAL ABSTRACT:** 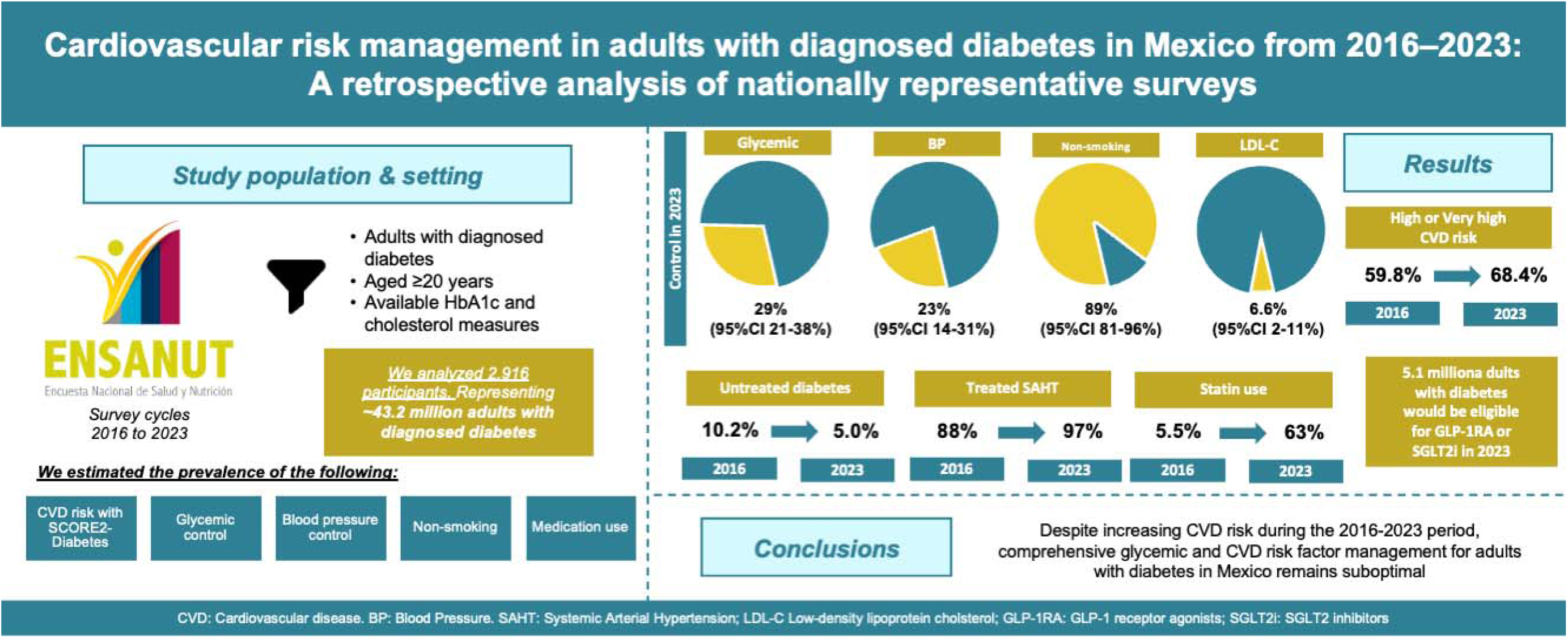

## INTRODUCTION

Diabetes mellitus, largely type 2 diabetes, is a growing health care problem in Mexico, with approximately 14.6 million individuals living with diabetes in 2022^1^. Diabetes is one of the leading causes of death in Mexico^2^, with at least one third of premature deaths attributable to this condition^3^, and significant excess risk associated with renal disease, cardiac disease, and stroke^4^. Cardiovascular disease (CVD) is the leading cause of mortality among people living with diabetes^5^, and common risk factors like hypertension, dyslipidemia, and smoking, which are particularly prevalent among individuals with diabetes in Mexico^6^, significantly contribute to the development of complications and increased risk of death^7,8^.

Consequently, clinical guidelines recommend multiple risk factor management in individuals with diabetes aimed at reducing risk of CVD beyond approaches solely focused on glycemic targets^5^. The introduction of novel glucose-lowering medications that can reduce major adverse CVD events is catalyzing progress in CVD-related mortality among people with diabetes, particularly in high income settings.^9,10^ However, comprehensive clinical diabetes management in middle-income countries like Mexico remains limited. The importance of strengthening CVD risk reduction among people with diabetes in Mexico is particularly important given that an estimated 30-40% of CVD deaths (cardiac, stroke, or vascular conditions) can be directly attributable to diabetes^3^. Furthermore, achievement of multicomponent CVD treatment goals in diabetes in Mexico has not been reported, with previous studies focusing solely on glycemic control^11,12^. Here, we aimed to: (1) estimate the prevalence of CVD risk categories using SCORE2-Diabetes, (2) assess the prevalence of glycemic, blood pressure (BP), cholesterol, non-current smoking, and combined risk factor control, and (3) explore trends in medication use and determinants of control achievement among individuals living with diabetes in Mexico.

## METHODS

### Study population

We analyzed data from the cross-sectional National Health and Nutrition Surveys (ENSANUT) cycles 2016, 2018, 2021, 2022, and 2023. Briefly, ENSANUT is a probabilistic survey which uses multistage, stratified, clustering sampling representative at national, regional, and rural/urban level^13–16^. In each cycle, individuals are interviewed, and information is collected regarding sociodemographic, lifestyle, and self-reported medical history. Clinical measurements (blood pressure, weight, and height) are obtained, and a 10 mL blood sample is collected from a random subsample to undergo subsequent biochemical analyses. Further details on ENSANUT’s methodology and data collection are reported elsewhere^13–16^. For this study, we included individuals with previously diagnosed diabetes aged ≥20 years, with available and complete HbA1c and cholesterol measures. We excluded individuals with undiagnosed diabetes, as our primary objective was to estimate achievement of clinical goals linked to medical treatment, which could only be ascertained in individuals with previous medical diagnosis of diabetes.

### Variable definitions

#### Diagnosed diabetes and glycemic control

Diagnosed diabetes was defined as self-reported medical diagnosis of diabetes. Glycemic control was defined based on age-specific clinical recommendations: HbA1c <7% for individuals aged <65 years and HbA1c <7.5% for those aged ≥65 years^17^.

#### Hypertension and BP control

BP was considered controlled if the mean measurement of systolic BP (SBP) and diastolic BP (DBP) were both <130/80 mmHg^5^. Diagnosed hypertension was defined as an individual with self-reported medical diagnosis of hypertension and undiagnosed hypertension as individuals without medical diagnosis but with SBP ≥140 mmHg or SBP ≥90 mmHg^18^. Hypertension treatment prevalence was estimated among individuals with diagnosed hypertension. Given that ENSANUT does not specify medications used by participants, we were unable to determine use of specific glucose or BP-lowering medications.

#### Cardiovascular risk categories, cholesterol control and statin treatment

CVD risk in individuals with diagnosed diabetes was assessed using SCORE2-Diabetes^19^ for a moderate-risk region (100–150 CVD deaths per 100,000 people), based on a reported rate of 146.3 CVD deaths per 100,000 people in Mexico in 2023^20,21^. SCORE2-Diabetes 10-year CVD risk categories were classified according to 2023 ESC Guidelines as low (<5%), moderate (5 to <10%), high (10 to <20%) and very high risk (SCORE2-Diabetes ≥20%, clinically established atherosclerotic CVD or severe target-organ damage)^18^. Due to the high prevalence of hypertriglyceridemia in the Mexican population, LDL-C was estimated using the Sampson equation^22,23^. LDL-C control was defined according to CVD risk categories as <55 mg/dL for very high, <70mg/dL for high and <100mg/dL for moderate and low CVD risk^18^. Eligibility for primary CVD prevention with statins was defined as any individual ≥40 years without previous CVD, and for secondary prevention as individuals ≥40 years with previous CVD^24^. We also estimated eligibility for GLP-1 receptor agonists and SGLT-2 inhibitors as first-line treatment based on SCORE2-Diabetes as per ESC guidelines^18^.

#### Non-current smoking and combined goal achievement

Non-current smoking was defined as individuals reporting being either never smokers or former smokers. To evaluate combined control achievement amongst individuals with diabetes in Mexico, we constructed composite variables according to whether participants achieved BP and LDL-C control (BC), glycemic, BP, and LDL-C control (ABC), or glycemic, BP, LDL-C, and non-current smoking (ABCN)^25^. Given our observation of low control achievement for LDL-C using SCORE2-Diabetes risk categories thresholds, combined control prevalence was estimated using less stringent LDL-C control targets based on ADA and Mexican guidelines as <100mg/dL in primary and <70mg/dL for sensitivity analyses.

#### Density-independent social lag index

To assess state-level social disadvantage we used the density-independent social lag index (DISLI) by obtaining residuals from a linear regression of population density onto SLI, which is a composite measure of access to education, health care, dwelling quality, and basic services in Mexico^26^.

### Statistical analyses

Weighted prevalence estimates were obtained considering complex survey design and sample weights from ENSANUT. We obtained prevalence estimates stratified by sex, age group (20-44, 45-64, ≥65 years), geographic location (urban or rural), indigenous identity (indigenous or non-indigenous), educational level (no education, elementary, middle/high school, university, other), and SCORE2-Diabetes risk categories (low/moderate and high/very high risk). The proportion of participants with missing data across pooled ENSANUT cycles ranged from 1.76% for HbA1c in 2022 to 12.4% for DBP in 2018. To determine the potential association between control achievement and several variables of interest, we fitted logistic regression models adjusted by sex, age group, educational level, social security affiliation (with or without social security), survey cycle, and DISLI category. All statistical analyses were conducted using R version 4.4.2, prevalence estimates were obtained using the *survey* R package, and a significance threshold of p<0.05 was used.

## RESULTS

### Characteristics of individuals with diagnosed diabetes

We analyzed data from 2,916 participants, representing over 43.2 million adults ≥20 years with diagnosed diabetes in Mexico during the 2016-2023 period. Individuals with diagnosed diabetes in Mexico were primarily women with a higher proportion of individuals in the age group of 45-64 years (**Supplementary Table 1**). Mean HbA1c levels had a downward trend from 2016 to 2018, with subsequent increments during the 2021-2023 period. Mean SBP ranged from 129 mmHg (95%CI 125–133 mmHg) in 2016 to 139 mmHg (95%CI 134–144 mmHg) in 2023, while mean DBP remained relatively steady across survey cycles. The prevalence of diagnosed hypertension increased from 42% (95%CI 35%-50%) in 2016 to 68% (95%CI 56%–78%) in 2023. Undiagnosed hypertension was stable across survey cycles, ranging from 10% (95%CI 6.6%-15%) in 2016 to 11% (95%CI 5.0%–21%) in 2023. Most individuals with diagnosed diabetes were eligible for primary prevention, accounting for 85% (95%CI 78%–90%) of individuals in 2016 and up to 93% (95%CI 85%-97%) in 2023. The proportion of individuals eligible for secondary prevention ranged from 4.1% (95%CI 2.7%–6.3%) in 2016 to 8.1% (95%CI 3.7%–17%) in 2021.

Important changes in social security affiliation (which in Mexico, provides access to healthcare, pensions, and other benefits, primarily based on formal employment status), were observed, with a high proportion of individuals insured during 2016-2018, and a decrease after 2021-2023. The highest share of participants with diagnosed diabetes was observed in urban settings and individuals with non-indigenous identity.

#### Glycemic control, BP control, and non-current smoking

Among individuals with diagnosed diabetes, glycemic control in Mexico during 2016-2023 remained suboptimal, varying from 36% (95%CI 28%–44%) in 2016 to 29% (95%CI 21%-38%) in 2023 (**Figure 1A**). BP control decreased from 45% (95%CI 37%–54%) in 2016 to 23% (95%CI 14%–31%) in 2023 (**Figure 1B**). Non-current smoking was high, with a modest increase in control across survey years, increasing from 86.4% (95%CI 79.0%– 93.9%) in 2016 to 88.7% (95%CI 81.2%–96.3%) in 2023 (**Figure 1C**).

**Figure 1.**
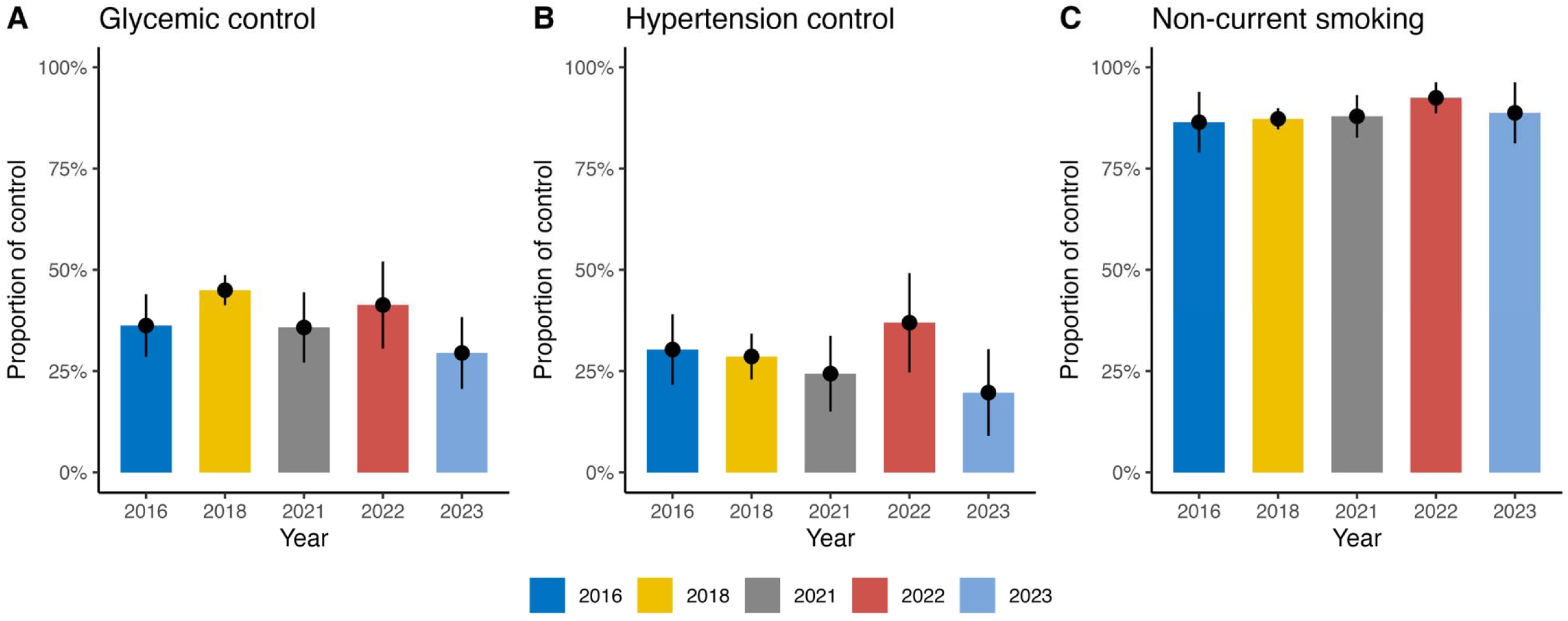
Prevalence of control achievement in individuals with diagnosed diabetes in Mexico during 2016-2023. (**A**) Prevalence of glycemic control, (**B**) prevalence of blood pressure control, (**C**) and prevalence of smoking control.

#### Prevalence of CVD risk and LDL-C control

The prevalence of high or very high 10-year CVD risk according to SCORE2-Diabetes amongst individuals with diagnosed diabetes increased from 59.8% (95%CI 52.1%-67.0%) in 2016 to 68.4% (95%CI 55.6%-78.9%) in 2023. Overall, high and very high CVD risk categories remained the most prevalent from 2016 to 2023. (**Figure 2A**). Most participants with diagnosed diabetes at high or very high risk received oral treatment in 2023, despite only 24.6% (95%CI 13.1%-36.2%) achieving glycemic control (**Figure 2B**). BP control was more adequate across CVD risk categories and high rates of BP-lowering treatment use were achieved in 2023 (**Figure 2C**). Regarding LDL-C control, among those at very-high risk, only 1.2% (95%CI 0.0%-3.3%) achieved the recommended LDL-C <50 mg/dL target. Similarly, only 0.4% (95%CI 0.0%-1.2%) of those at high risk achieved the LDL-C <70mg/dL goal, and for those at moderate and low CVD risk, 26.2% (95%CI 2.1-50.3%) and 20.3% (95%CI 1.4%-39.3%) achieved LDL-C <100 mg/dL, respectively (**Figure 2D**). Over time, LDL-C control based on SCORE2-Diabetes risk categories showed a slight increase ranging from 2.8% (95%CI 1.2%-4.4%) in 2016 to 6.6% (95%CI 1.9%-11.2%) in 2023. Statin use was suboptimal regardless of CVD risk categories (**Figure 2E**). In 2023, amongst those at very-high or high risk, a statin was used by 69.8% (95%CI 38.6%-100%), and amongst those at low or moderate risk, in 53.3% (95%CI 25.6%-81.1%) of participants. Finally, we estimated the total number of individuals that would be eligible to receive GLP-1 receptor agonists or SGLT2 inhibitors in 2023, which would be 5,097,988 (95%CI 3,096,065-7,159,911) adults ≥20 years in Mexico with diagnosed diabetes.

**Figure 2.**
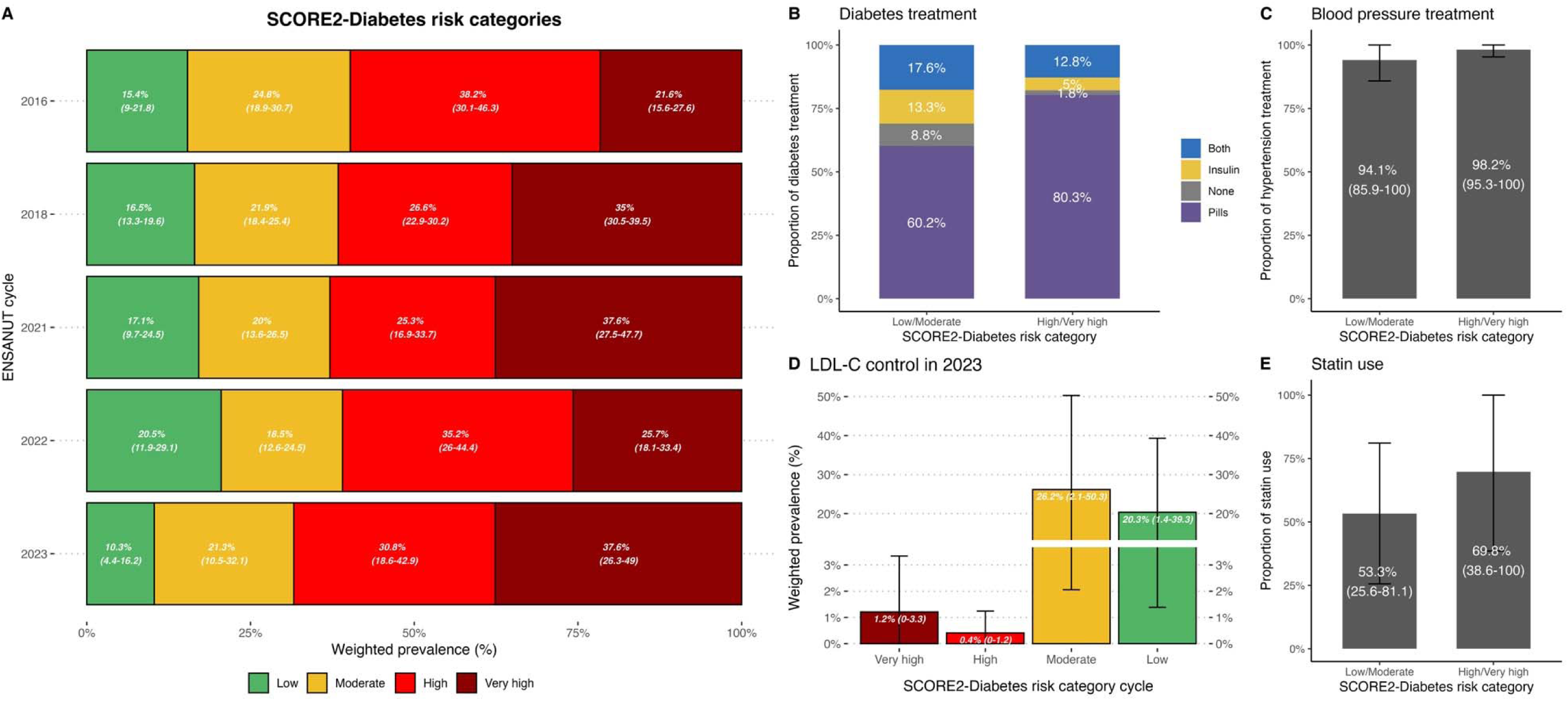
Prevalence of cardiovascular disease risk categories using SCORE2-Diabetes amongst individuals with diagnosed diabetes in Mexico during 2016-2023 (**A**). The figure also shows the prevalence of diabetes-related treatment modalities (**B**), blood pressure lowering treatment (**C)**, LDL-C goal achievement according to SCORE2-Diabetes categories (**D)** and statin use (**E**), comparing adults with diagnosed diabetes at low/moderate compared to high/very high risk of cardiovascular disease using SCORE2-Diabetes. SCORE2-Diabetes 10-year CVD risk categories were classified according to 2023 ESC Guidelines as low (<5%), moderate (5 to <10%), high (10 to <20%) and very high risk (SCORE2-Diabetes ≥20%, clinically established atherosclerotic CVD or severe target-organ damage).

### Prevalence of combined control achievement in individuals with diagnosed diabetes

Combined achievement was low, primarily due to suboptimal LDL-C control, even using less stringent goals. Using LDL-C <70 mg/dL as the target, BC control ranged from 0.12% (95%CI 0.0%–0.34%) in 2016 to 1.03% (95%CI 0.0%-2.3%) in 2022 and ABCN control ranged from 0.11% (95%CI 0.0%–0.34%) in 2016 to 1.5% (95%CI 0.0%–3.1%) in 2023 (**Supplementary Figure 1**). Using <100 mg/dL as LDL-C target combined control improved, with BC control ranging from 2.6% (95%CI 0.9%–4.3%) in 2016 to 7.2% (95%CI 2.5%–12.0%) in 2023. Similarly, using this definition, ABCN control increased from 1.4% (95%CI 0.1%–2.5%) in 2016 to 4.6% (95%CI 2.1%–7.1%) in 2022 (**Supplementary Figure 1).**

### Modifiers of control achievement in individuals with diagnosed diabetes

When stratifying by sex, glycemic control prevalence was similar among men and women, and BP control prevalence was higher among women. Interestingly, LDL-C control was suboptimal for both men and women. While men achieved slightly higher rates of control than women, non-current smoking was higher in women (**Supplementary Figure 3**). BC control prevalence was higher in men. Stratifying by age, individuals ≥65 years had a higher prevalence of glycemic and non-current smoking compared to other groups. However, individuals 20-44 years had higher prevalence of BP and LDL control (**Supplementary Figure 4**). Stratified analyses by geographic location show slightly better LDL-C control in rural areas and by indigenous identity better glycemic control among individuals with non-indigenous identity (**Supplementary Figure 5** and **Supplementary Figure 6)**.

### Medication use amongst individuals with diagnosed diabetes

Treatment with oral glucose-lowering medications remained stable over time, with increases in combined insulin and oral treatment modalities (**Figure 3A**). The prevalence of untreated individuals decreased from 10.2% (95%CI 5.8%–14.7%) in 2016 to 5.0% (95%CI 0.7%–9.3%) in 2023. Amongst individuals with diagnosed hypertension, the proportion of those with BP-lowering treatment was high, ranging from 88% (95%CI 81%– 95%) in 2016 to 97% (95%CI 93%–99%) in 2023 (**Figure 3B**). Of note, overall statin use increased significantly across survey cycles, from 5.5% (95%CI 2.5%–8.5%) in 2016, 12% (95%CI 9.1%–14%) in 2018, 51% (95%CI 38%–63%) in 2021, 58% (95%CI 43%–73%) in 2022, and 63% (95%CI 41.1%-85.2%) in 2023 (**Figure 3C**), which is consistent with increases in LDL-C control over time. Amongst individuals eligible for primary prevention according to ADA guidelines, statin use increased from 5.9% (95%CI 2.5%–9.4%) in 2016 to 64% (95%CI 41%–87%) in 2023. Statin use amongst individuals eligible for secondary prevention showed a similar albeit smaller increase from 7.3% (95%CI 0%–18%) in 2016 to 28% (95%CI 0%–84%) in 2022. There was an unusual increase in 2021, with a prevalence of statin use of 73% (95%CI 39%–100%) in this group, although these estimations should be used with caution due to the small number of individuals in this subset (**Supplementary Figure 6**).

**Figure 3.**
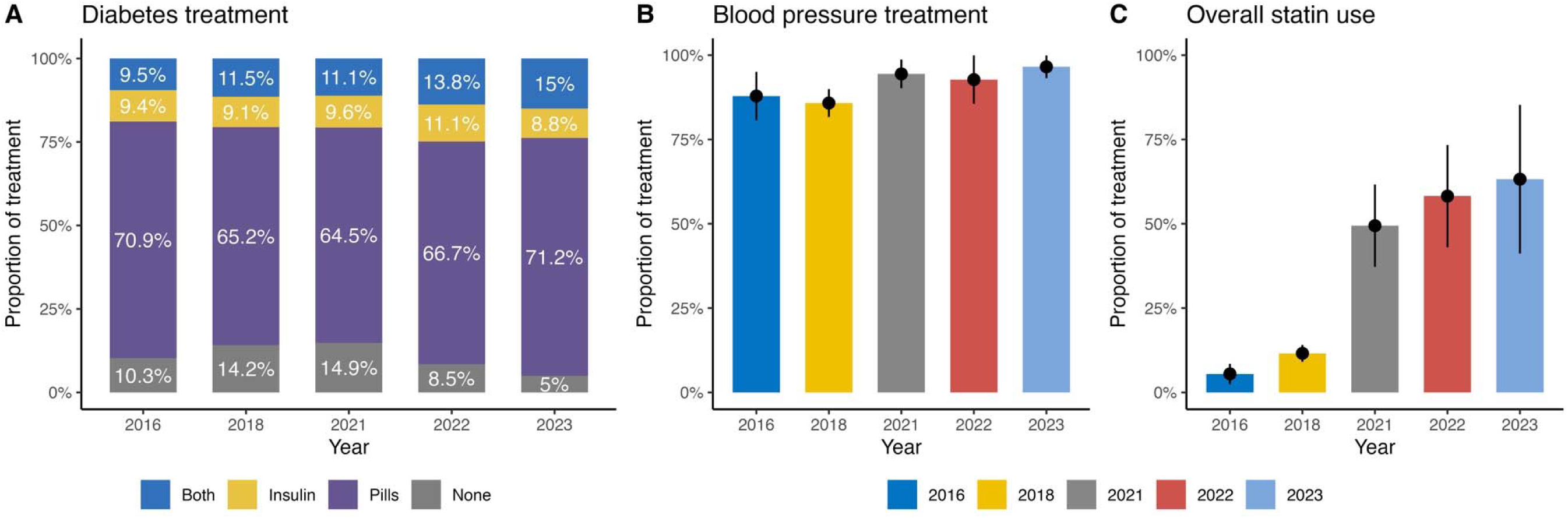
Prevalence of medication use amongst individuals with diagnosed diabetes during the 2016-2023 period. (**A**) Prevalence of diabetes medication use amongst individuals with diagnosed diabetes, (**B**) Prevalence of antihypertensive medication use amongst individuals with diagnosed hypertension, (**C**) Prevalence of overall statin use.

### Determinants of control achievement

Compared to women, men had higher odds of achieving glycemic and LDL-C control, but lower odds of achieving BP and non-current smoking. Age groups 40-59 years and ≥60 years had lower odds of achieving glycemic, BP and LDL control and of having low/moderate CVD risk compared to individuals aged 20–44 years. However, these older age groups had higher odds of achieving non-current smoking (**Table 1**). Compared to individuals without education, those with college education had higher odds of achieving glycemic and BP control, without consistent associations for LDL-C or non-current smoking. Lower DISLI categories were associated with higher odds of achieving glycemic control compared to higher DISLI, but a negative association was found for BP, LDL, low/moderate CVD risk, and non-current smoking. Additionally, individuals in the survey cycle 2023 had much higher odds of achieving LDL-C control compared to participants in the 2016 cycle. Finally, participants with social security had lower odds of achieving control and of having low/moderate CVD risk, but higher odds of achieving BP, and non-current smoking compared to those without social security affiliation. Finally, women and individuals in the age group 20–44 years had higher odds of achieving ABCN control, with lower odds among those who were in the low DISLI categories.

**TABLE 1.** Multivariate logistic regression estimates evaluating the adjusted odds ratio (aOR) of achieving glycemic, blood pressure, cholesterol (<70 mg/dL), smoking, BC, and ABCN control in individuals with diagnosed diabetes in Mexico. 95% CI: 95% confidence interval.

|  | Glycemic control |  | BP control |  | Cholesterol control |  | Smoking control |  | Low or Moderate CVD risk |  | BC control |  | ABCN control |  |
| --- | --- | --- | --- | --- | --- | --- | --- | --- | --- | --- | --- | --- | --- | --- |
| Characteristic | aOR | 95% CI | aOR | 95% CI | aOR | 95% CI | aOR | 95% CI | aOR | 95% CI | aOR | 95% CI | aOR | 95% CI |
| **Sex (Ref. Women)** |  |  |  |  |  |  |  |  |  |  |  |  |  |  |
| *Men* | 1.17 | 1.09, 1.24 | 0.49 | 0.47, 0.51 | 1.46 | 1.38, 1.55 | 0.20 | 0.19, 0.21 | 0.95 | 0.88, 1.02 | 0.66 | 0.60, 0.72 | 0.38 | 0.36, 0.41 |
| **Age group (Ref. <40 years)** |  |  |  |  |  |  |  |  |  |  |  |  |  |  |
| *40-59 years* | 0.23 | 0.21, 0.25 | 0.45 | 0.43, 0.47 | 0.70 | 0.66, 0.75 | 1.48 | 1.40, 1.57 | 0.51 | 0.47, 0.56 | 0.47 | 0.42, 0.52 | 0.26 | 0.23, 0.29 |
| *≥60 years* | 0.18 | 0.17, 0.20 | 0.28 | 0.26, 0.30 | 0.65 | 0.60, 0.71 | 2.41 | 2.22, 2.62 | 0.42 | 0.37, 0.47 | 0.45 | 0.39, 0.52 | 0.01 | 0.01, 0.01 |
| **Educational level (Ref. No education)** |  |  |  |  |  |  |  |  |  |  |  |  |  |  |
| *Elementary school* | 0.77 | 0.68, 0.86 | 1.05 | 0.96, 1.14 | 0.86 | 0.76, 0.97 | 1.00 | 0.88, 1.14 | 0.68 | 0.57, 0.81 | 0.75 | 0.61, 0.92 | 1.57 | 1.41, 1.73 |
| *Middle/High school* | 1.03 | 0.91, 1.18 | 1.21 | 1.10, 1.32 | 0.88 | 0.78, 1.01 | 1.00 | 0.88, 1.14 | 0.86 | 0.72, 1.03 | 0.97 | 0.79, 1.18 | 2.24 | 2.00, 2.51 |
| *University* | 1.34 | 1.15, 1.57 | 1.29 | 1.17, 1.42 | 0.68 | 0.59, 0.79 | 1.17 | 1.02, 1.34 | 0.63 | 0.52, 0.77 | 0.71 | 0.57, 0.89 | 2.95 | 2.55, 3.41 |
| *Other* | 1.09 | 0.90, 1.32 | 1.24 | 1.10, 1.40 | 0.38 | 0.30, 0.47 | 1.32 | 1.10, 1.58 | 0.37 | 0.27, 0.49 | 0.33 | 0.23, 0.48 | 1.49 | 1.26, 1.78 |
| **Indigenous identify (Ref. Non-indigenous)** |  |  |  |  |  |  |  |  |  |  |  |  |  |  |
| *Indigenous* | 0.80 | 0.71, 0.89 | 1.08 | 1.00, 1.16 | 1.32 | 1.18, 1.46 | 2.33 | 2.06, 2.64 | 1.18 | 1.02, 1.36 | 1.41 | 1.20, 1.64 | 1.18 | 1.05, 1.34 |
| **Survey cycle (Ref. year 2016)** |  |  |  |  |  |  |  |  |  |  |  |  |  |  |
| *2018* | 1.02 | 0.92, 1.14 | 0.72 | 0.67, 0.77 | 1.13 | 1.01, 1.27 | 1.00 | 0.92, 1.09 | 0.91 | 0.78, 1.07 | 0.79 | 0.66, 0.94 | 1.06 | 0.95, 1.18 |
| *2021* | 0.89 | 0.78, 1.02 | 0.89 | 0.82, 0.96 | 1.57 | 1.37, 1.79 | 1.02 | 0.92, 1.13 | 1.66 | 1.40, 1.99 | 1.68 | 1.38, 2.05 | 0.84 | 0.74, 0.96 |
| *2022* | 0.71 | 0.62, 0.82 | 0.87 | 0.80, 0.94 | 6.76 | 5.97, 7.67 | 0.83 | 0.75, 0.92 | 6.85 | 5.83, 8.09 | 5.43 | 4.53, 6.57 | 0.57 | 0.49, 0.65 |
| *2023* | 0.70 | 0.59, 0.83 | 1.06 | 0.93, 1.21 | 5.57 | 4.82, 6.45 | 0.83 | 0.73, 0.96 | 4.62 | 3.80, 5.63 | 3.20 | 2.55, 4.03 | 1.59 | 1.31, 1.94 |
| **DISLI category (Ref. DISLI High)** |  |  |  |  |  |  |  |  |  |  |  |  |  |  |
| *Low* | 1.14 | 1.06, 1.24 | 0.87 | 0.83, 0.92 | 0.93 | 0.86, 0.99 | 0.98 | 0.92, 1.05 | 0.64 | 0.58, 0.70 | 0.66 | 0.59, 0.73 | 1.44 | 1.32, 1.57 |
| *Moderate* | 1.20 | 1.10, 1.31 | 0.89 | 0.85, 0.94 | 0.83 | 0.77, 0.90 | 0.78 | 0.73, 0.83 | 0.68 | 0.61, 0.75 | 0.66 | 0.59, 0.74 | 1.01 | 0.93, 1.11 |
| *Very Low* | 1.20 | 1.08, 1.34 | 0.89 | 0.83, 0.94 | 0.80 | 0.73, 0.87 | 0.66 | 0.61, 0.71 | 0.68 | 0.61, 0.77 | 0.60 | 0.52, 0.68 | 1.00 | 0.90, 1.11 |
| **Social security (Ref. without social security)** |  |  |  |  |  |  |  |  |  |  |  |  |  |  |
| Characteristic | aOR | 95% CI | aOR | 95% CI | aOR | 95% CI | aOR | 95% CI | aOR | 95% CI | aOR | 95% CI | aOR | 95% CI |
| *With social security* | 0.81 | 0.74, 0.88 | 1.09 | 1.04, 1.14 | 0.98 | 0.92, 1.05 | 1.10 | 1.04, 1.17 | 1.04 | 0.96, 1.13 | 0.99 | 0.90, 1.09 | 0.84 | 0.77, 0.92 |

## DISCUSSION

Comprehensive management of CVD risk amongst adults with diagnosed diabetes in Mexico remains suboptimal. Over two thirds of adults with diagnosed diabetes in Mexico have high or very high 10-year CVD risk using SCORE2-Diabetes and most of them do not achieve CVD control goals. Our estimates show that less than half of individuals with diagnosed diabetes achieve glycemic or BP control, even though a high proportion of them report medication use. Furthermore, LDL-C control is poor in Mexican adults with diagnosed diabetes in whom, despite a consistent increase in statin use across survey cycles, stringent LDL-C goals are seldom met, irrespective of any specific guideline threshold. Additionally, although non-current smoking increased steadily throughout the study period, combined target control was rarely achieved, primarily due to inadequate cholesterol control. Interestingly, our results suggest that over 5.1 million adults with diagnosed diabetes in Mexico may be eligible to receive GLP-1 receptor agonists and SGLT-2 inhibitors as first-line pharmacotherapy, as most have high or very high predicted 10-year CVD risk. Our results are consistent with previous evidence which suggests some progress in diabetes management in Mexico, primarily related to glycemic and non-current smoking, and more recently on statin adoption. However, these improvements are still insufficient, and comprehensive management strategies are underutilized or not widely implemented^27,28^.

Diabetes is the second leading cause of death in Mexico^2^, and has been associated with mortality rates twice as high as those observed in high income countries^4^. Given that disease prevalence has increased significantly in the last two decades^1,29^, suboptimal control of diabetes carries profound implications for public health in Mexico, leading to disparities in the diabetes presentation and related complications^30,31^. The failure to achieve CVD targets is probably related to multiple causes, including lack of widespread use of new generation glucose-lowering medications in Mexico (which might also be influenced by affordability issues), limited access to high-quality diabetes care, physician therapeutic inertia, and a generalized lack of disease awareness. These results highlight the need to address, not only individual-level factors (i.e. improving patient education or physician training), but broader structural issues that might hinder CVD target achievement (i.e. fragmented medical care or socioeconomic inequalities).

Uncontrolled risk factors are associated with higher risk of complications and death among individuals with diabetes, and combined interventions aimed at controlling risk factors have shown to reduce micro- and macrovascular complications, and mortality^7,32,33^. Accordingly, recent clinical guidelines emphasize a comprehensive approach to reduce CVD risk factors giving special priority to management of glycemia, BP, and lipids^5^. Previous studies in the Mexican population have reported increasing disease prevalence and poor glycemic control throughout the last two decades^30^. In 2006, only 3.5% of individuals with diabetes in Mexico were within glycemic control targets, and in 2012 this proportion increased substantially to 29%^29,34^. Our estimates show that there was a continued, albeit moderate improvement in glycemic control over the first half of the past decade, reaching 36% in 2016, and 45% in 2018. This trend, however, stalled and declined after the COVID-19 pandemic, which was associated with important disruptions to Mexican health services^35^, and a significant increase in diabetes-related excess mortality in Mexico^27^. Therefore, we hypothesize that the observed reduction in glycemic control observed in our study might be directly related to limited access to medical care during the pandemic. Interestingly, pharmacologic therapy for glycemic control has been high for the last two decades, with previous studies consistently finding more than 80% of individuals with diagnosed diabetes reporting medication use^12,34^. Our estimates are in line with these results, with every survey cycle analyzed presenting more than 84% of diabetes medication adoption. These findings suggest that suboptimal glycemic control might be related to lack of access to quality medical care but also to lack of disease awareness, therapeutic inertia, or lack of physician compliance regarding clinical recommendations, all of which should be explored and addressed to improve diabetes management in Mexico. Additionally, costs and availability of novel glucose-lowering medications should be strongly considered to optimize diabetes care and reduce CVD risk.

BP control estimates remained suboptimal for the 2016-2023 period, despite more than 85% of individuals with diagnosed hypertension reporting use of antihypertensive medication. A previous analysis of the Mexico City Prospective Study (MCPS) found much smaller proportions of antihypertensive use, changing from 35% in 1998-2004 to 51% in 2015-2019, with an associated increase in the adoption of angiotensin-converting enzyme inhibitors (ACEI) and angiotensin II receptor blockers (ARB)^36^. Although these estimations are lower than those found in ENSANUT, it should be noted that MCPS is not a representative cohort, and recruitment was only done in urban participants from Mexico City. To our knowledge there are no other nationally representative studies exploring BP control in individuals with diabetes in Mexico, so temporal trends of control are limited to our estimations. Notably, our analysis found that one out of six individuals with diagnosed diabetes in Mexico had undiagnosed hypertension. Given that hypertension in diabetes is related to increased risk of CVD events, heart failure, and microvascular complications^37^, and the fact that Mexican and international clinical guidelines recommend BP measurements at every clinical visit^5,38^, increasing awareness of comorbid hypertension amongst individuals with diabetes represents a significant area of improvement for the Mexican healthcare system, which could have direct implications on reducing the burden of diabetes on Mexican individuals.

Our results showed that LDL-C targets were the least likely to be achieved, with our estimates suggesting a low prevalence of control from 2016 to 2023, regardless of target definition and CVD risk category. This finding is particularly worrisome given that reducing LDL-C levels has been strongly associated with reductions in CVD events, vascular mortality, and all-cause mortality^39,40^. Recent clinical recommendations have advocated for stringent LDL-C targets, emphasizing high intensity statin therapy in combination with ezetimibe or PCSK9 inhibitor to reach these goals^5^. In Mexico, however, statin adoption has been slow and affordability of other lipid lowering medications is a concern; notably, no previous studies have analyzed cholesterol management in Mexican individuals with diabetes. A previous analysis of the MCPS found that <1% of individuals with diagnosed diabetes reported using lipid-lowering medications in 1998-2004, with a slight increase to 14% in 2015-2019^36^. Our results show that in 2016 overall statin use was low, with a prevalence of 5.6%, which then showed a significant increase to 64% in 2023. Even though this increase is an important step in cholesterol management, statin adoption appears to be insufficient, given that only one in four individuals with diabetes achieved an LDL-C level of <100 mg/dL by 2023 and even fewer achieved a level of LDL-C <70 mg/dL or LDL-C control according to the more stringent ESC guidelines. Additionally, our results show that statin use may still be insufficient for individuals eligible for secondary prevention, which carry the highest risk and the bigger potential benefit. Of relevance, the statin intensity or the use of specific statins could not be evaluated in our study and should be further explored to identify areas of opportunity to improve prescription patterns for statins to improve cholesterol management for individuals with diabetes in Mexico.

Our study has several strengths. To our knowledge, this is the first study to evaluate the comprehensive CVD risk management of diabetes in Mexico, while most have focused primarily on prevalence and glycemic control. The use of a series of nationally representative surveys allowed to obtain population-level estimates of diabetes control, and the availability of information since 2016 facilitates describing the most recent population-level trends. There are also some limitations that should be considered to adequately interpret our results. First, the small sample size of individuals with diagnosed diabetes in each survey cycle might reduce the accuracy of estimated and reduce statistical power to detect small but meaningful changes over time. Second, we were unable to determine the type of glucose-lowering, BP-lowering and lipid-lowering medications used by participants, as well as their prescribed doses and adherence, which represents important elements related to quality and accessibility of diabetes management. Third, whilst SCORE2-Diabetes may be a useful tool to improve assessment of CVD risk in diabetes, its use has not been validated in Mexican population and its consistent use in clinical practice will require further validation studies. Finally, the cross-sectional nature of our analysis does not allow us to ascertain causal relationships in the observed associations, particularly for determinants of treatment goal achievements.

In conclusion, achievement of treatment goals in individuals with diagnosed diabetes remained suboptimal in Mexico during the 2016-2023 period. Over two thirds of adults with diagnosed diabetes have high or very high predicted CVD risk in 2023 and most do not reach adequate control targets. Specifically, achievement of glycemic and BP control was observed in less than half of adults with diagnosed diabetes, non-current smoking improved significantly, whilst LDL-C control was low (regardless of LDL-C goals), even with a high proportion of participants reporting consistent medication use. Our results suggest a need to improve comprehensive CVD risk factor management in individuals with diabetes beyond glycemic control, which is essential to improve diabetes care and reduce the risk of diabetes-related mortality and complications in Mexico^27,30^.

## Data Availability

DATA AVAILABILITY: All code and materials are available for reproducibility of results at https://github.com/oyaxbell/diabetescare_ensanut/

https://ensanut.insp.mx/index.php

## ACKNOWLEDGMENTS

This project was registered and approved by the Research Committee at Instituto Nacional de Geriatría, project number DI-PI-009-2024. JPE and CAFM are enrolled at the PECEM Program of the Faculty of Medicine at UNAM and are supported by CONACyT. JAS was supported by Grant Number K23DK135798 from the NIH/NIDDK and by the Massachusetts General Hospital Executive Committee and Center for Diversity and Inclusion Physician-Scientist Development Award.

## AUTHOR CONTRIBUTIONS

Research idea and study design: DRG, PNML, JS, OYBC; data acquisition: CAFM, DRG, PNML, OYBC; analysis/interpretation: DRG, PNML, OYBC; statistical analysis: DRG, PNML, OYBC; manuscript drafting: DRG, PNML, CAFM, JPE, JPDS, CDPC, AVV, MTLT, NEAV, GD, JAS, OYBC; supervision or mentorship: OYBC. Each author contributed important intellectual content during manuscript drafting or revision and accepts accountability for the overall work by ensuring that questions pertaining to the accuracy or integrity of any portion of the work are appropriately investigated and resolved.

## DATA AVAILABILITY

All code and materials are available for reproducibility of results at https://github.com/oyaxbell/diabetescare_ensanut/

## ETHICAL DISCLOSURES

This project was registered and approved by the Research Committee at Instituto Nacional de Geriatría, project number DI-PI-009-2024.

