## Supplementary Material for "Cardiovascular risk management in adults with diagnosed diabetes in Mexico from 2016–2023: A retrospective analysis of nationally representative surveys"

**SUPPLEMENTARY TABLES**

| Characteristic | ENSANUT 2016<br>n=454<br>N = 6,453,990<br>(95% CI)[12] | ENSANUT 2018<br>n=1,630<br>N = 8,500,126<br>(95% CI)[12] | ENSANUT 2021<br>n=304<br>N = 8,376,509<br>(95% CI)[12] | ENSANUT 2022<br>n=313<br>N = 10,460,069<br>(95% CI)[12] | ENSANUT 2023<br>n=215<br>N = 9,438,145<br>(95% CI)[12] |
|---|---|---|---|---|---|
| Sex (%) | | | | | |
| Women | 61 (53%, 69%) | 60 (56%, 64%) | 52 (43%, 61%) | 59 (50%, 67%) | 56 (45%, 67%) |
| Men | 39 (31%, 47%) | 40 (36%, 44%) | 48 (39%, 57%) | 41 (33%, 50%) | 44 (33%, 55%) |
| Age group (%) | | | | | |
| <40 years | 11 (6.1%, 18%) | 7.8 (6.0%, 10%) | 7.3 (3.4%, 15%) | 10 (5.5%, 18%) | 5.8 (2.1%, 15%) |
| 40-59 years | 47 (39%, 55%) | 44 (40%, 48%) | 42 (33%, 52%) | 41 (31%, 52%) | 36 (26%, 47%) |
| ≥60 years | 43 (35%, 51%) | 48 (44%, 52%) | 50 (41%, 60%) | 48 (38%, 59%) | 58 (47%, 68%) |
| HbA1c (%) | 8.44 (8.0, 8.9) | 7.98 (7.8, 8.2) | 8.38 (8.0, 8.8) | 8.12 (7.7, 8.6) | 8.88 (8.2, 9.5) |
| SBP (mmHg) | 129 (125, 133) | 135 (133, 137) | 133 (130, 137) | 130 (126, 134) | 139 (134, 144) |
| DBP (mmHg) | 75 (73, 77) | 76 (75, 77) | 78 (76, 80) | 74 (72, 75) | 79 (74, 83) |

| Characteristic | ENSANUT 2016<br>n=454<br>N = 6,453,990<br>(95% CI)[1,2] | ENSANUT 2018<br>n=1,630<br>N = 8,500,126<br>(95% CI)[1,2] | ENSANUT 2021<br>n=304<br>N = 8,376,509<br>(95% CI)[1,2] | ENSANUT 2022<br>n=313<br>N = 10,460,069<br>(95% CI)[1,2] | ENSANUT 2023<br>n=215<br>N = 9,438,145<br>(95% CI)[1,2] |
|---|---|---|---|---|---|
| Diabetes treatment (%) | | | | | |
| None | 10 (6.6%, 16%) | 14 (12%, 17%) | 15 (8.8%, 24%) | 8.5 (5.0%, 14%) | 5.0 (2.1%, 12%) |
| Pills | 71 (63%, 77%) | 65 (61%, 69%) | 64 (55%, 73%) | 67 (57%, 75%) | 71 (61%, 79%) |
| Insulin | 9.4 (5.4%, 16%) | 9.1 (6.8%, 12%) | 9.6 (5.9%, 15%) | 11 (7.4%, 16%) | 8.8 (4.6%, 16%) |
| Both | 9.5 (6.6%, 13%) | 11 (9.3%, 14%) | 11 (6.5%, 18%) | 14 (6.9%, 26%) | 15 (8.5%, 25%) |
| Hypertension treatment (%) | | | | | |
| Untreated | 12 (6.5%, 21%) | 14 (11%, 19%) | 5.6 (2.5%, 12%) | 7.3 (2.6%, 19%) | 3.5 (1.3%, 9.0%) |
| Treated | 88 (79%, 93%) | 86 (81%, 89%) | 94 (88%, 97%) | 93 (81%, 97%) | 97 (91%, 99%) |
| Education level (%) | | | | | |

| Characteristic | ENSANUT 2016<br>n=454<br>N = 6,453,990<br>(95% CI)[12] | ENSANUT 2018<br>n=1,630<br>N = 8,500,126<br>(95% CI)[12] | ENSANUT 2021<br>n=304<br>N = 8,376,509<br>(95% CI)[12] | ENSANUT 2022<br>n=313<br>N = 10,460,069<br>(95% CI)[12] | ENSANUT 2023<br>n=215<br>N = 9,438,145<br>(95% CI)[12] |
|---|---|---|---|---|---|
| No education | 14 (10%, 19%) | 11 (9.1%, 14%) | 12 (7.2%, 20%) | 11 (6.8%, 17%) | 5.4 (2.9%, 9.6%) |
| Elementary school | 42 (36%, 49%) | 44 (40%, 48%) | 38 (30%, 47%) | 30 (23%, 38%) | 39 (29%, 51%) |
| Middle/High school | 32 (24%, 41%) | 31 (28%, 35%) | 34 (26%, 44%) | 41 (32%, 52%) | 36 (26%, 47%) |
| University | 6.6 (3.6%, 12%) | 8.8 (6.6%, 12%) | 13 (7.1%, 22%) | 12 (6.5%, 21%) | 15 (7.8%, 27%) |
| Other | 5.2 (2.5%, 11%) | 4.9 (3.5%, 7.0%) | 2.6 (1.3%, 5.0%) | 5.9 (2.6%, 13%) | 4.6 (1.6%, 12%) |
| Social security (%) | | | | | |
| Without social security | 7.3 (4.2%, 12%) | 7.7 (6.0%, 10%) | 36 (27%, 45%) | 31 (23%, 40%) | 35 (25%, 46%) |
| With social security | 93 (88%, 96%) | 92 (90%, 94%) | 64 (55%, 73%) | 69 (60%, 77%) | 65 (54%, 75%) |
| Location (%) | | | | | |
| Rural | 22 (18%, 28%) | 17 (15%, 20%) | 16 (11%, 23%) | 16 (11%, 23%) | 25 (13%, 42%) |

| Characteristic | ENSANUT 2016<br>n=454<br>N = 6,453,990<br>(95% CI)[12] | ENSANUT 2018<br>n=1,630<br>N = 8,500,126<br>(95% CI)[12] | ENSANUT 2021<br>n=304<br>N = 8,376,509<br>(95% CI)[12] | ENSANUT 2022<br>n=313<br>N = 10,460,069<br>(95% CI)[12] | ENSANUT 2023<br>n=215<br>N = 9,438,145<br>(95% CI)[12] |
|---|---|---|---|---|---|
| Urban | 78 (72%, 82%) | 83 (80%, 85%) | 84 (77%, 89%) | 84 (77%, 89%) | 75 (58%, 87%) |
| Indigenous identity (%) | | | | | |
| Non-indigenous | 90 (80%, 95%) | 94 (92%, 95%) | 91 (82%, 96%) | 95 (89%, 97%) | 98 (94%, 99%) |
| Indigenous | 9.9 (4.6%, 20%) | 6.2 (4.5%, 8.3%) | 9.0 (4.3%, 18%) | 5.4 (2.7%, 11%) | 2.0 (0.67%, 6.0%) |
| Hypertension status (%) | | | | | |
| Without hypertension | 48 (40%, 56%) | 34 (31%, 38%) | 41 (32%, 51%) | 45 (36%, 53%) | 22 (13%, 33%) |

| Characteristic | ENSANUT 2016<br>n=454<br>N = 6,453,990<br>(95% CI)[12] | ENSANUT 2018<br>n=1,630<br>N = 8,500,126<br>(95% CI)[12] | ENSANUT 2021<br>n=304<br>N = 8,376,509<br>(95% CI)[12] | ENSANUT 2022<br>n=313<br>N = 10,460,069<br>(95% CI)[12] | ENSANUT 2023<br>n=215<br>N = 9,438,145<br>(95% CI)[12] |
|---|---|---|---|---|---|
| Diagnosed hypertension | 42 (35%, 50%) | 50 (46%, 54%) | 46 (37%, 56%) | 43 (34%, 52%) | 68 (56%, 78%) |
| Undiagnosed hypertension | 10 (6.6%, 15%) | 15 (12%, 19%) | 13 (7.1%, 21%) | 13 (7.0%, 22%) | 11 (5.0%, 21%) |
| CV risk prevention (%) | | | | | |
| Not eligible for CV risk prevention (<40 years) | 11 (6.1%, 18%) | 7.8 (6.0%, 10%) | 7.4 (3.4%, 15%) | 10 (5.5%, 18%) | 5.8 (2.1%, 15%) |
| Eligible for primary prevention | 85 (78%, 90%) | 85 (82%, 87%) | 85 (75%, 91%) | 85 (76%, 92%) | 93 (85%, 97%) |

| Characteristic | ENSANUT 2016<br>n=454<br>N = 6,453,990<br>(95% CI)[1,2] | ENSANUT 2018<br>n=1,630<br>N = 8,500,126<br>(95% CI)[1,2] | ENSANUT 2021<br>n=304<br>N = 8,376,509<br>(95% CI)[1,2] | ENSANUT 2022<br>n=313<br>N = 10,460,069<br>(95% CI)[1,2] | ENSANUT 2023<br>n=215<br>N = 9,438,145<br>(95% CI)[1,2] |
|---|---|---|---|---|---|
| Eligible for secondary prevention | 4.1 (2.7%, 6.3%) | 7.5 (5.7%, 10%) | 8.1 (3.7%, 17%) | 4.4 (1.3%, 14%) | 1.2 (0.46%, 3.0%) |

[1]%; Mean

[2]CI = Confidence Interval

HbA1c: Glycated hemoglobin, SBP: Systolic blood pressure, DBP: Diastolic blood pressure.

**Suppementary Table 1.** Weighted characteristics of interviewed individuals with diagnosed diabetes according to ENSANUT survey cycle. Each survey shows the total number of participants with diabetes along with the expanded population of individuals with diabetes to whom these estimates are representative using ENSANUT complex survey design.

### SUPPLEMENTARY FIGURES

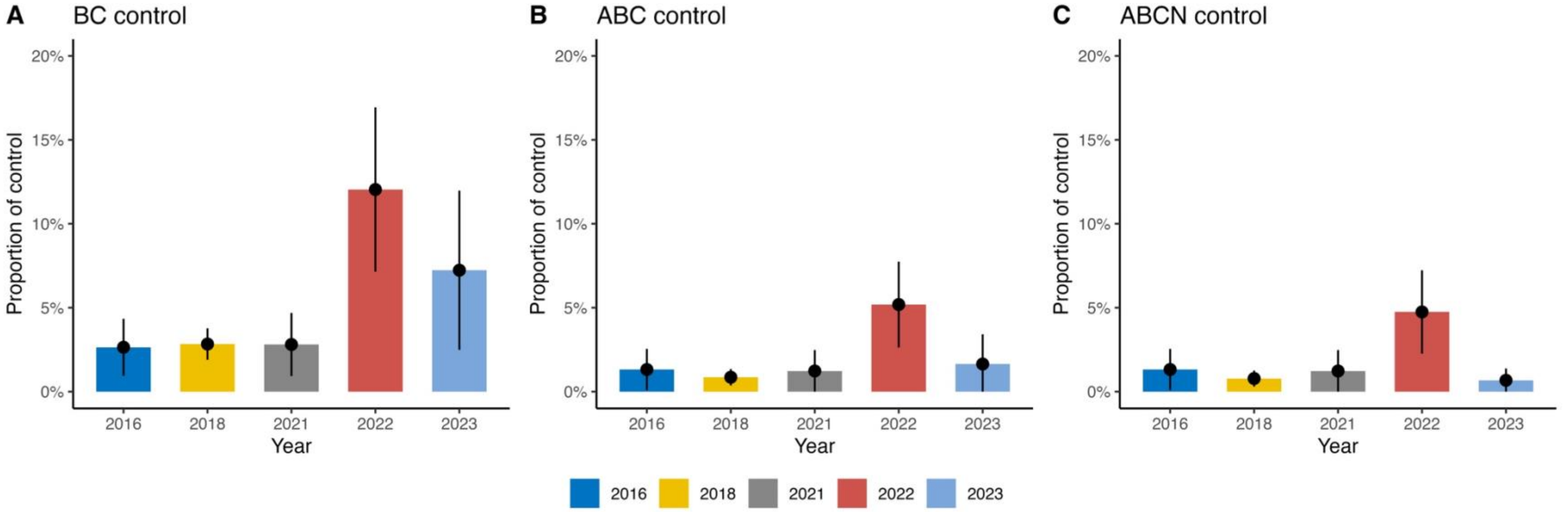


**Supplementary Figure 1.** Prevalence of combined control achievement in individuals with diagnosed diabetes in Mexico during 2016-2023, defined as blood pressure and LDL-C control (BC), glycemic, blood pressure, and LDL-C control (ABC), or glycemic, blood pressure, LDL-C, and smoking control (ABCN) using LDL-C targets <100mg/dL.

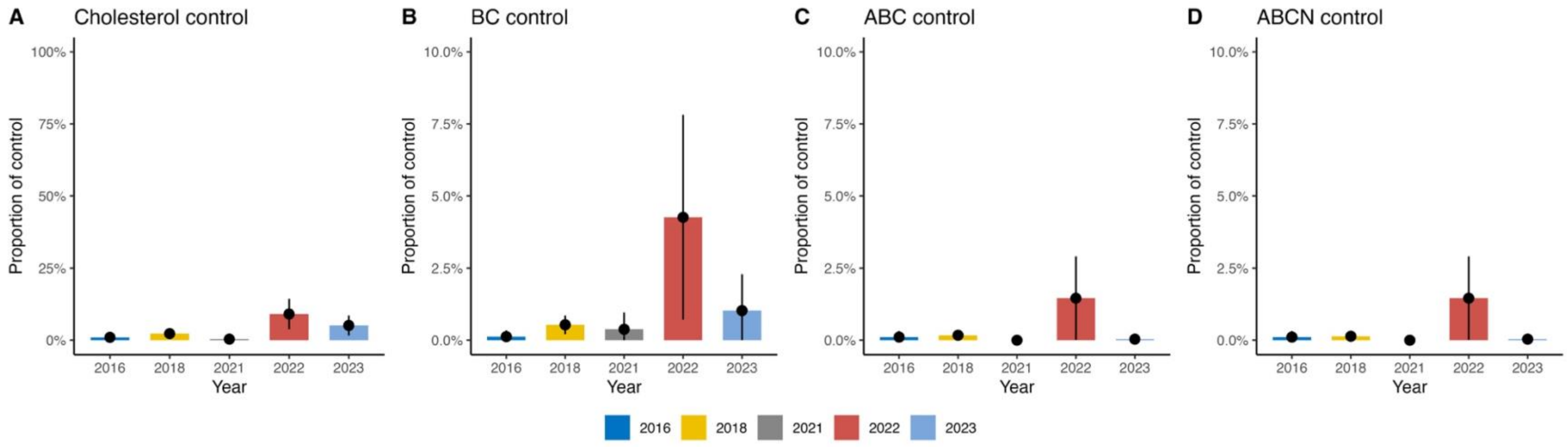


**Supplementary Figure 2.** Prevalence of combined control achievement in individuals with diagnosed diabetes in Mexico during 2016-2023, defined as blood pressure and LDL-C control (BC), glycemic, blood pressure, and LDL-C control (ABC), or glycemic, blood pressure, LDL-C, and smoking control (ABCN) using LDL-C targets <70mg/dL.

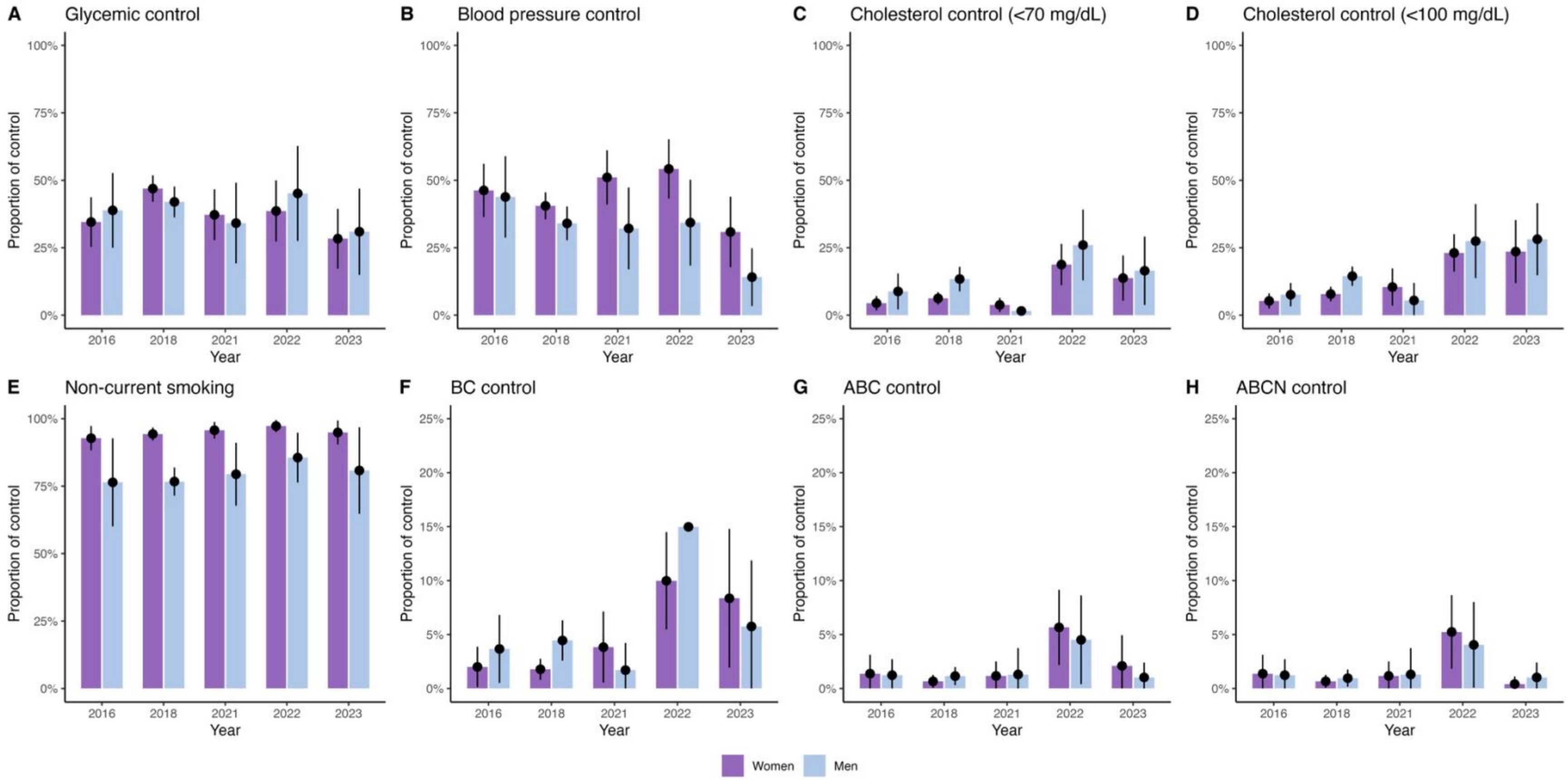


**Supplementary Figure 3.** Prevalence of control achievement in individuals with diagnosed diabetes in Mexico during 2016-2023 stratified by sex. (A) Prevalence of glycemic control, (B) Prevalence of blood pressure control, (C) Prevalence of cholesterol control (<70 mg/dL), (D) Prevalence of cholesterol control (<100 mg/dL), (E) Prevalence of smoking control.

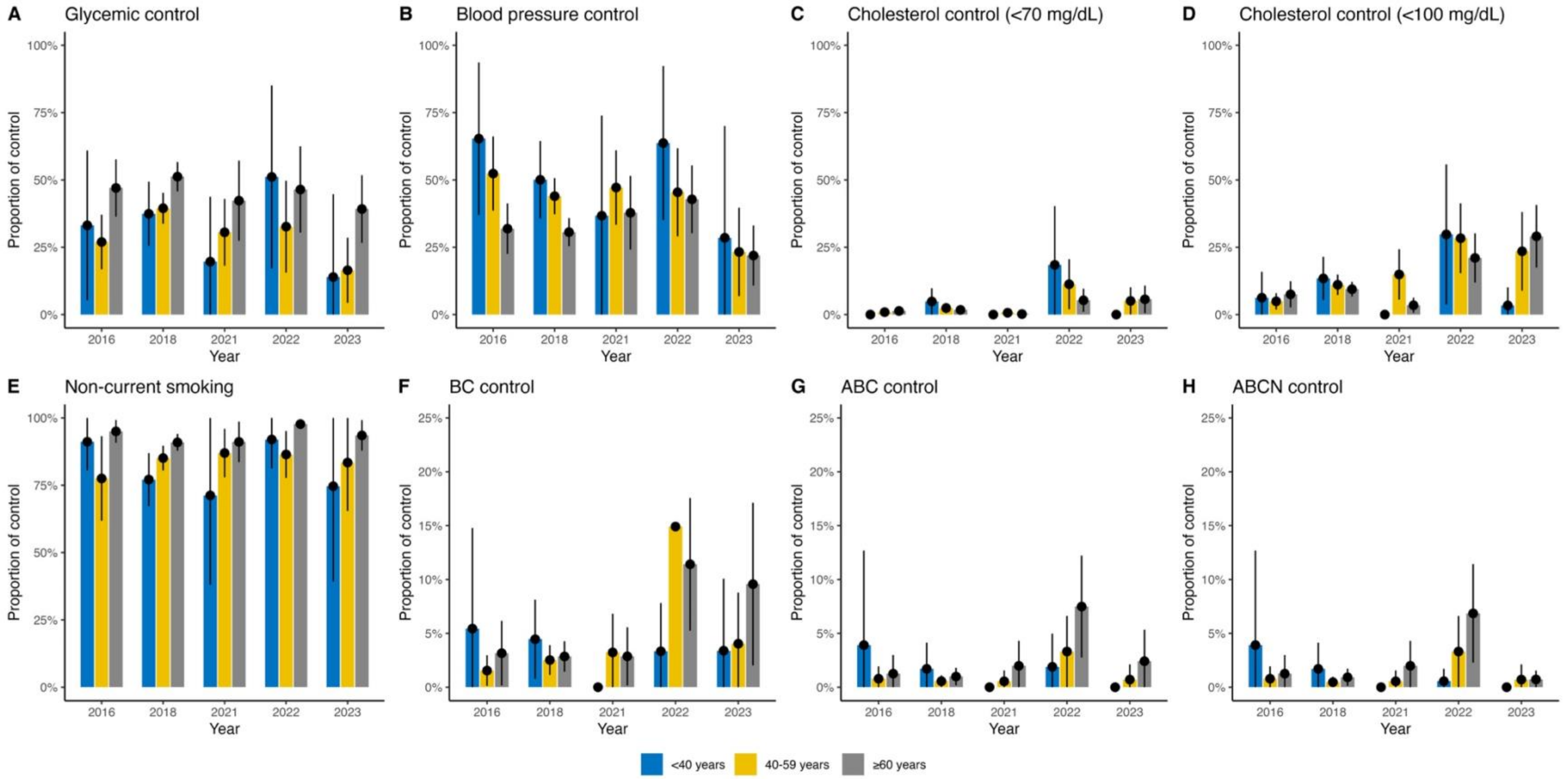


**Supplementary Figure 4.** Prevalence of control achievement in individuals with diagnosed diabetes in Mexico during 2016-2023 stratified by age groups. (A) Prevalence of glycemic control, (B) Prevalence of blood pressure control, (C) Prevalence of cholesterol control (<70 mg/dL), (D) Prevalence of cholesterol control (<100 mg/dL), (E) Prevalence of smoking control.

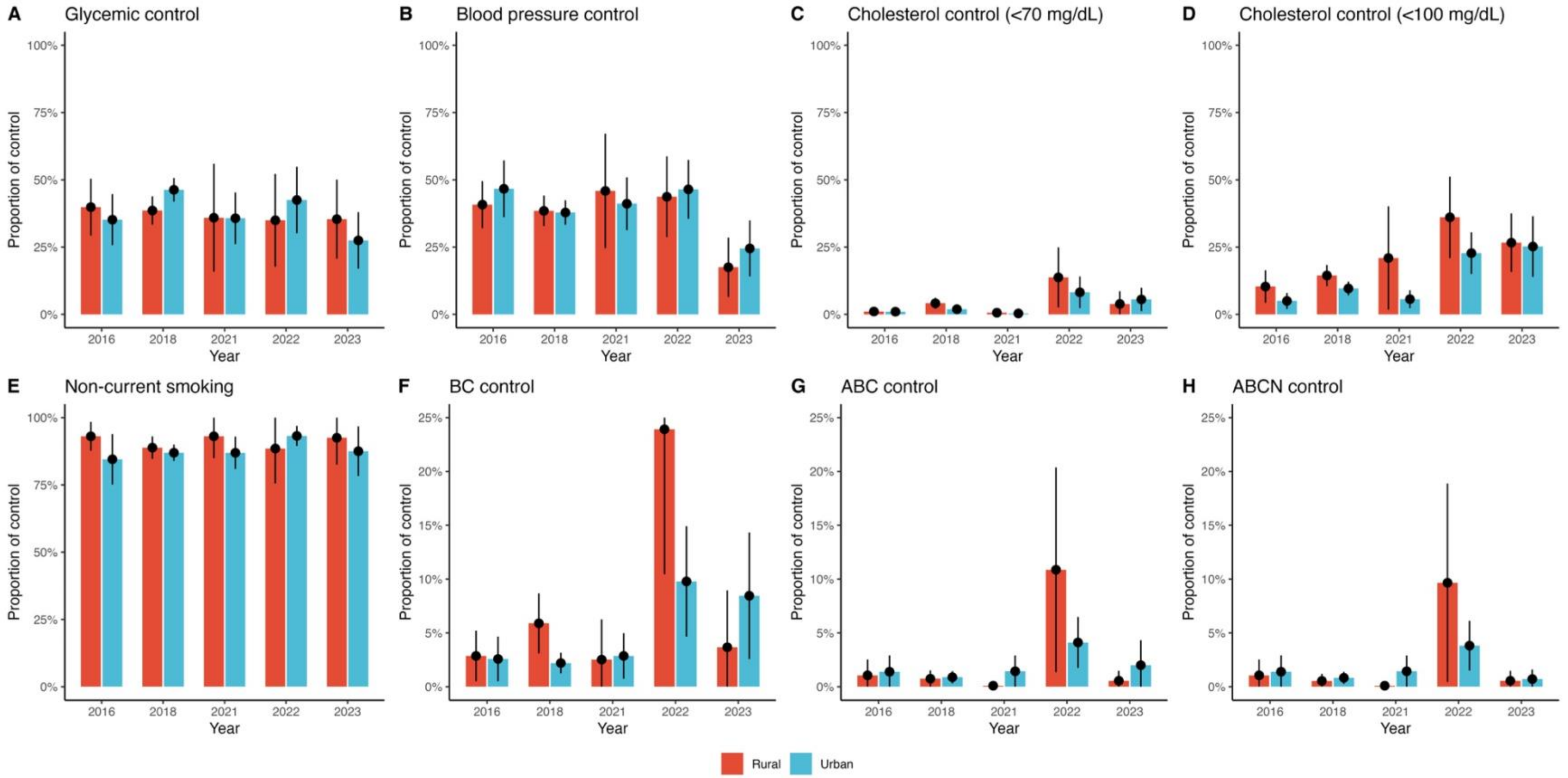


**Supplementary Figure 5.** Prevalence of control achievement in individuals with diagnosed diabetes in Mexico during 2016-2023 stratified by urban versus rural region. (A) Prevalence of glycemic control, (B) Prevalence of blood pressure control, (C) Prevalence of cholesterol control (<70 mg/dL), (D) Prevalence of cholesterol control (<100 mg/dL), (E) Prevalence of smoking control.

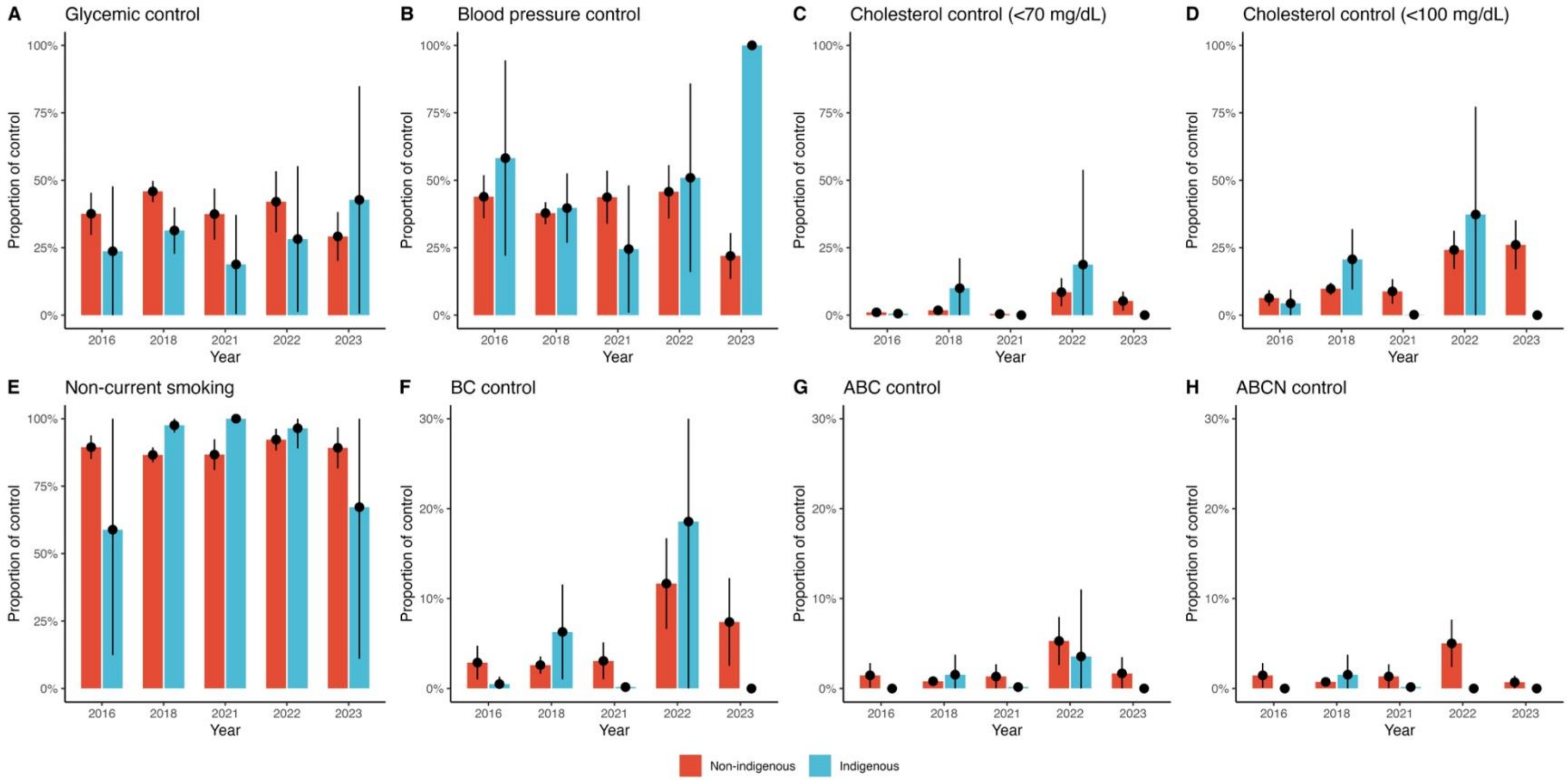


**Supplementary Figure 6.** Prevalence of control achievement in individuals with diagnosed diabetes in Mexico during 2016-2023 stratified by indigenous identity. (A) Prevalence of glycemic control, (B) Prevalence of blood pressure control, (C) Prevalence of cholesterol control (<70 mg/dL), (D) Prevalence of cholesterol control (<100 mg/dL), (E) Prevalence of smoking control.

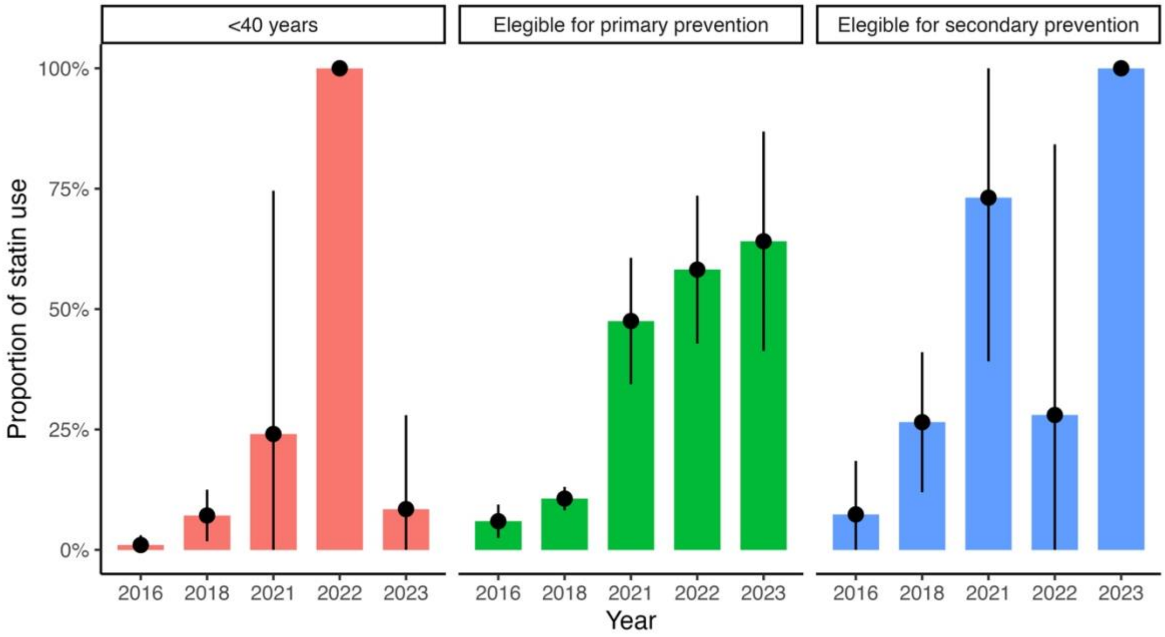


**Supplementary Figure 7.** Prevalence of statin users amongst individuals with diabetes eligible for primary and secondary prevention during the 2016-2023 period.
